# Predicting Bacterial Vaginosis Development using Artificial Neural Networks

**DOI:** 10.1101/2025.05.02.25326872

**Authors:** Jacob H. Elnaggar, John W. Lammons, Caleb M. Ardizzone, Kristal J. Aaron, Clayton Jacobs, Keonte J. Graves, Sheridan D. George, Meng Luo, Ashutosh Tamhane, Paweł Łaniewski, Alison J. Quayle, Melissa M. Herbst-Kralovetz, Nuno Cerca, Christina A. Muzny, Christopher M. Taylor

## Abstract

Bacterial vaginosis (BV) is a dysbiosis of the vaginal microbiome, characterized by the depletion of protective *Lactobacillus* spp. and overgrowth of anaerobes. Artificial neural network (ANN) modeling of vaginal microbial communities offers an opportunity for early detection of incident BV (iBV). 16S rRNA gene sequencing and quantitative PCR was performed on longitudinal vaginal specimens collected from participants within 14 days of iBV or from healthy participants to calculate the inferred absolute abundance (IAA) of vaginal bacterial taxa. ANNs were trained using the IAA of vaginal taxa from 420 vaginal specimens to classify individual vaginal specimens as either pre-iBV (collected before iBV onset) or Healthy. Feature importance was assessed to understand how specific vaginal micro-organisms contributed to model predictions. ANN modeling accurately classified >97% of specimens as either pre-iBV or Healthy (sensitivity >96%, specificity >98%) using IAA of 20 vaginal taxa. Model prediction accuracy was maintained when training models using only a few key vaginal taxa. Models trained using only the top five most important features achieved an accuracy of >97%, sensitivity >92%, and specificity >99%. Model predictive accuracy was further improved by training models on specimens from white and black participants separately; using only three feature models achieved an accuracy >96%, sensitivity >91%, and specificity >91%. Feature analysis found that *Lactobacillus* species *L. gasseri* and *L. jensenii* differed in how they contributed to model predictions in models trained with data stratified by race. A total of 420 vaginal specimens were analyzed, providing a robust dataset for model training and validation.

**Importance:** Bacterial vaginosis (BV) is the most common vaginal infection and is associated with numerous comorbidities. BV is associated with infertility, preterm birth, pelvic inflammatory disease, and increased risk of HIV/STI acquisition. BV is difficult to detect prior to onset, and infection commonly recurs after treatment. Our model allows for the accurate early detection of iBV by surveying the vaginal microbiome, potentially serving as a valuable tool to determine which patients are at risk of developing iBV. Early detection of iBV could lead to wider adoption of clinical interventions useful in the prevention of iBV such as live biotherapeutics, prophylactic antibiotics, and/or behavioral modifications. Our findings indicate that few microbial targets are required for accurate predictions, facilitating cost and time effective clinical testing. Similarly, our study highlights the value of developing models personalized to specific patient populations, improving accuracy while reducing the number of taxa required for accurate predictions.

## INTRODUCTION

Bacterial vaginosis (BV) is the most common vaginal infection, affecting approximately 30% of reproductive-age women worldwide.^1–3^ BV is characterized as a vaginal dysbiosis and is associated with multiple adverse health outcomes including infertility, adverse birth outcomes, increased risk of HIV and acquisition of sexually transmitted infections (STIs), pelvic inflammatory disease, and increased risk of post-gynecologic surgery pelvic infections.^1,4,5^ A large body of data suggest that BV is sexually transmitted,^6–8^ however, the inciting pathogen(s) remain controversial. It is unknown whether BV is caused by acquisition of a single pathogen, a polymicrobial consortium of pathogenic bacteria,^2,9–11^ or depletion of protective lactobacilli in the vaginal microbiota ^12–15^, which paves the way for colonization and infection with facultative and strictly anaerobic BV-associated bacteria. Lack of a clear understanding of BV pathogenesis has directly impeded advancements in diagnosis, treatment, and prevention of recurrent infection.^16–18^

Demographic factors such as race add to the complexity of BV pathogenesis considering vaginal microbiome composition and BV incidence differ significantly based on race.^19^ Developing novel diagnostics that account for race may aid in improving BV diagnosis and add clarity to the microbial features contributing to BV acquisition.

Application of machine learning to vaginal microbiome data presents an opportunity to advance BV research.^20^ An artificial neural network (ANN) is one such supervised machine learning tool, designed to identify complex patterns in data. An ANN consists of an interconnected, feed-forward network of input, hidden, and output neurons.^21,22^ In our application of ANNs for vaginal microbiome analysis, sequencing-generated quantitative data serve as inputs, with individual neurons representing distinct bacterial taxonomic classifications.^20^ These machine learning techniques can facilitate the investigation of the complexities of the vaginal microbiota by processing and interpreting vast amounts of microbial data. By applying ANNs to vaginal microbiome studies, analysis of subtle changes in the microbiome can occur, allowing further investigation of changes associated with incident BV (iBV). This approach enables the identification of specific microbial signatures and patterns that may be critical in understanding and diagnosing BV.

In this study, we applied ANNs to predict the development of iBV using data from 420 vaginal specimens collected in a prospective study.^23^ While the parent study tracked changes in vaginal microbial communities over time, our specialized ANN models allowed us to accurately predict whether a participant would develop iBV within the next two weeks based on a single vaginal specimen. We developed highly accurate models that assessed key vaginal microbial features, to personalize BV diagnosis and better characterize the microbial factors contributing to iBV pathogenesis.

## METHODS

### Clinical Enrollment

In a prospective, longitudinal study, we enrolled an ethnically diverse group of heterosexual women at the University of Alabama at Birmingham (UAB) Sexual Health Research Clinic in Birmingham, AL (UAB Institutional Review Board Protocol #300004547). Enrollment criteria are fully described in the published protocol paper.^23^ In brief, women included were English-speaking, between 18-45 years of age, and had a current male sexual partner. Those who had used oral or intravaginal antibiotics within the past 14 days, had self-reported HIV infection, or were currently pregnant were excluded. Women enrolled were asymptomatic with no Amsel criteria and a normal Nugent score of 0-3 with no *Gardnerella* morphotypes noted on baseline vaginal Gram stain.^24,25^ Participants were screened for baseline STIs (*Trichomonas vaginalis*, *Chlamydia trachomatis*, *Neisseria gonorrhoeae,* and *Mycoplasma genitalium*) by nucleic acid amplification testing and those testing positive were treated, per current recommendations,^26^ and dropped from the study.

The study design is similar to our previous iBV pathogenesis study;^27^ enrolled participants self-collected three vaginal specimens twice daily for 60 days. All specimens were collected with nylon-tipped flocked swabs (Copan Diagnostics) and placed in separate tubes of a 1:10 dilution of AssayAssure Sample Preservative (Sierra Molecular Corporation) and 1X phosphate buffered saline (PBS) solution. One of these vaginal swabs was used to smear a glass slide for future Nugent score determination in our research laboratory prior to placing it in a specimen collection tube. Women also completed daily diaries in which sexual activity, partner characteristics, vaginal symptoms, menses, douching activity, and oral/intra-vaginal medication use were recorded. All vaginal specimens were collected at home; collection tubes were refrigerated and vaginal smears were stored at room temperature in individual plastic slide holders to prevent breakage. All materials were delivered weekly to the study site for Gram stain and Nugent score determination (vaginal smears) or preservation at - 80°C (specimen collection tubes) until further testing.

### Selection of vaginal specimens for microbiome characterization

Participants were followed for iBV development throughout the duration of the study (Nugent score of 7-10 on ≥4 consecutive vaginal specimens). For women who developed iBV, twice daily vaginal specimens were selected from the 14 days prior to iBV (30 specimens total), the day of iBV, as well as three days post diagnosis of iBV (six specimens total).^25^ Healthy controls (who maintained normal vaginal microbiota for the majority of the study) were matched to iBV cases by age, race, contraceptive method, and day of menses; an equivalent number of vaginal specimens from healthy controls were selected. All vaginal specimens were shipped to the Microbial Genomics Resource Group (MGRG) at the Louisiana State University Health Sciences Center (LSUHSC) in New Orleans, LA for sequencing and analysis. The MGRG isolated DNA from vaginal specimens, as described previously, using a modified QIAamp DNA Mini Kit (QIAGEN).^27,28^

### Molecular Methods

As represented in **Figure 1A**, the isolated DNA from twice-daily vaginal specimens was used to perform 16S ribosomal RNA (rRNA) gene sequencing on vaginal specimens from women who developed iBV and matched healthy controls to determine changes in the relative abundance of vaginal bacteria over time. Quantitative PCR (qPCR) for the universal 16S rRNA gene was used to determine bacterial burden and calculate inferred absolute abundance (IAA) concentrations of vaginal organisms, as outlined in Tettamanti Boshier, et al.^29^

**Figure 1.**
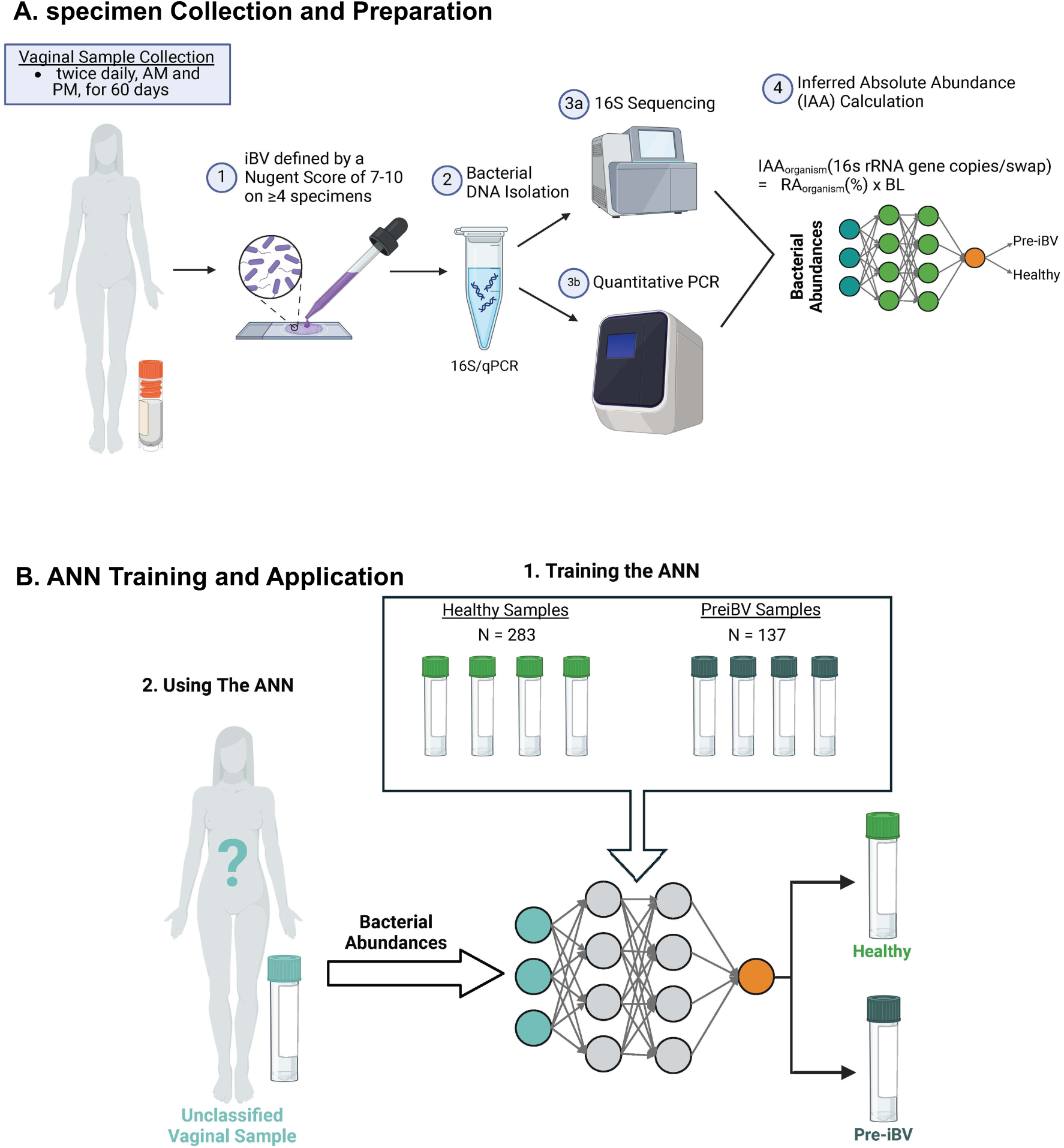
Study Workflow for Training ANNs to Predict preiBV from Vaginal specimens. (A) Twice daily vaginal specimens were collected from 16 participants and monitored for development of iBV using Nugent scoring. DNA was isolated from vaginal specimens which was used for 16s sequencing and qPCR assays to quantify vaginal taxa and bacterial load (BL). IAA of vaginal taxa was calculated from 16s relative abundance and bacterial loads. IAA abundance of key vaginal taxa was utilized to train ANN models to classify specimens as preiBV or Healthy. (B) ANN models were trained using IAA abundance of key vaginal taxa from individual specimens (n = 420) to predict ifa single specimen is from a participant that develops iBV within 14 days (preiBV) or does not develop iBV within the next 14 days (Healthy). The model is designed so that it can be used to classify a single vaginal specimen as Healthy or preiBV.

As in our previous study,^27^ the MGRG at LSUHSC performed 16S sequencing on the vaginal specimens. First, PCR amplicon libraries spanning the taxonomically informative fourth hypervariable (V4) region of the 16S rRNA were generated and sequenced on the Illumina MiSeq platform.^28^ Sequencing data were analyzed in R v4.2.1 using DADA2 v1.16,^30^ with taxonomic classification using SILVA v138.^31^ Further species classification of the *Lactobacillus* genus were performed using curated BLAST queries.^32^ We used the *Gardnerella*:*Prevotella*:*L.iners* mix standard, as described previously, to calculate the total bacterial load via broad range 16S rRNA gene qPCR within the vaginal specimens.^33^ We calculated the IAA of vaginal bacteria in our specimens using the following calculation: IAA (16S rRNA gene copies/specimen) = Relative Abundance (%) × Total Bacterial Load (universal 16S rRNA gene copies/specimen).^29^

### Modeling approach and parameters

An ANN is a type of statistical model similar to nonlinear regression models.^34^ Input for the model was the abundance of vaginal taxa as measured through IAA. We used specimens from iBV cases and healthy controls as input. Using the sequencing data obtained from the twice daily vaginal specimens, we built and compared several ANNs using the software packages TensorFlow (v2.16.1) and Keras (v3.0).^35,36^ First, we grouped the iBV specimens and healthy control specimens into two groupings: “pre-iBV” from the days leading up to the onset of iBV and “Healthy” from the women who did not develop iBV. The data were then split, in which 80% were inputted into the network with the labels iBV and Healthy to train the ANN. The remaining 20% was used for validation, without these labels, to assess how well the network was trained. Additionally, each ANN was trained over 600 iterations (epochs) of the training-and- validation process. We compared several architectures, combinations of hidden layers, to achieve the most optimal accuracy. This range of input structures and model architectures is known to be optimal for similar omics datasets.^37^ Stochastic gradient descent was used as the optimization algorithm, and neurons used a Rectified Linear Unit (ReLU) activation function. Furthermore, a grid search approach was used to determine optimal number of hidden layers, node combinations, dropout percentages, batch size, and regularization to maximize the accuracy of predicting influential BV-associated bacteria in the development of iBV. We also experimented with alternative optimizers and activation functions. Final models were selected based on training and validation accuracy. ANNs were implemented in Python3 in the visual studio code (VS code v1.96.2). Visualizations were created using matplotlib v3.7.1.^38^

## Determination of key BV-associated bacteria in the development of iBV

We used a mixed model neural network that combined mixed models with ANNs to allow for random effects in repeated sampling of the same participant.^39^ Once the ANN was trained and validated, we deconstructed the model using Shapely additive explanations (SHAP) (v0.46.0) Kernel Explainer method,^40^ to determine the “relative importance” of features included in each model. The data used to train models was included as the background while testing data were used to compute SHAP values for individual predictions. For models trained on 100 or more specimens, SHAP explainer background was limited to 100 training specimens. Mean absolute SHAP values were calculated for each feature by averaging SHAP values across all specimens to assess feature importance for each model. Additionally, SHAP values provided the opportunity to assess how features contributed to model predictions for individual specimens. Mean absolute SHAP values were used to select features for each model trained on a subset of features.

### Sample size estimation

Sample size was determined based on convenience sampling. All specimens available at the time of analysis were included in the study.

### Statistical analysis

Participant characteristics were statistically compared using a t-test for numerical covariates, and a chi-square test or Fisher’s exact test for categorical covariates. A p-value of 0.05 determined statistical significance and all analysis were conducted using SAS 9.4 (Cary, NC).

### Replicates

All qPCR reactions were initially performed in duplicate. To assess the consistency between technical replicates, we compared the cycle threshold (CT) values and considered a difference of ≤0.5 CT between wells as acceptable, given the logarithmic nature of qPCR quantification. If the CT variation exceeded this threshold—indicating potential technical error—the reaction was repeated in triplicate to resolve discrepancies and ensure data reliability. While duplicates carry some risk of undetected outliers, this stepwise validation minimized technical variability and reinforced the robustness of our quantification process.

### Randomization/Blinding

Not Applicable

### Ethics Statement

This study was approved by the University of Alabama at Birmingham Institutional Review Board (UAB IRB Protocol #300004547) with a Reliance Agreement at LSUHSC New Orleans. Written informed consent was obtained for all participants prior to enrollment in the study. All specimens and participant data were de-identified prior to use in this project.

## RESULTS

### Clinical sampling and modeling workflow

For this study, we analyzed data from 420 vaginal specimens that were self-collected by participants, encompassing both pre-iBV and healthy specimens.^23^ **Table 1** displays the demographic data for all participants from which vaginal specimens were used for this study. Features known to alter the vaginal microbiome such as age, race, education, STI history, and contraception use did not differ significantly between participants in the study who contributed vaginal specimens to the pre-iBV and Healthy groups (p <0.05).

**Table 1.** Characteristics of iBV Case and Control Participants at Study Enrollment. *STIs include chlamydia, gonorrhea, trichomoniasis, genital herpes, and human papilloma virus (HPV). ^#^Includes birth control pills, hormonal IUDs, and implants. ^$^Includes copper IUDs. Does not include protective barriers, such as condoms, diaphragms, or spermicides. ^&^History of contraception use does not include current contraception users. Abbreviations: SD = standard deviation, BV=bacterial vaginosis, STI=sexually transmitted infection; IUD=intrauterine device

| Characteristic | Total N=16 | iBV cases<br>(n=8) | Controls<br>(n=8) | p-value |
| --- | --- | --- | --- | --- |
| **Age (years ± SD)** | 29.2 ± 8.4 | 29.8 ± 8.6 | 28.6 ± 8.7 | 0.798 |
| **Race (self-identified)** |  |  |  | 0.285 |
| White | 8 (50.0) | 3 (37.5) | 5 (62.5) |  |
| African American | 6 (37.5) | 3 (37.5) | 3 (37.5) |  |
| Asian | 2 (12.5) | 2 (25.0) | 0 (0.0) |  |
| **Ethnicity (self-identified)** |  |  |  | 1.00 |
| Hispanic | 1 (6.3) | 0 (0.0) | 1 (12.5) |  |
| Non-Hispanic | 15 (93.8) | 8 (100.0) | 7 (87.5) |  |
| **Education** |  |  |  | 0.219 |
| Any post-graduate studies | 5 (31.2) | 4 (50.0) | 1 (12.5) |  |
| Bachelor's degree | 3 (18.8) | 1 (12.5) | 2 (25.0) |  |
| Some college/Associate's degree | 7 (43.8) | 2 (25.0) | 5 (62.5) |  |
| High school/GED | 1 (6.2) | 1 (12.5) | 0 (0.0) |  |
| Less than high school | 0 (0.0) | 0 (0.0) | 0 (0.0) |  |
| **Current Smoker** |  |  |  |  |
| Yes | 0 (0.0) | 0 (0.0) | 0 (0.0) |  |
| No | 16 (100.0) | 8 (100.0) | 8 (100.0) |  |
| **STI History\*** |  |  |  | 0.84 |
| *≥2 STIs* | 4 (25.0) | 2 (25.0) | 2 (25.0) |  |
| *1 STI* | 5 (31.2) | 3 (37.5) | 2 (25.0) |  |
| *None* | 7 (43.8) | 3 (37.5) | 4 (50.0) |  |
| **History of BV** |  |  |  | 1.00 |
| *Yes* | 6 (37.5) | 3 (37.5) | 3 (37.5) |  |
| *No* | 10 (62.5) | 5 (62.5) | 5 (62.5) |  |
| **Current Contraception Use** |  |  |  | 0.91 |
| *Hormonal Contraception<sup>#</sup>* | 10 (62.5) | 5 (62.5) | 5 (62.5) |  |
| *Birth Control Pills* | 3 (18.8) | 2 (25.0) | 1 (12.5) |  |
| *Hormonal IUD* | 5 (31.2) | 2 (25.0) | 3 (37.5) |  |
| *Implant* | 2 (12.5) | 1 (12.5) | 1 (12.5) |  |
| *Non-Hormonal Contraception<sup>\$</sup>* | | | | |
| *Copper IUD* | 0 (0.0) | 0 (0.0) | 0 (0.0) |  |
| *None* | 6 (37.5) | 3 (37.5) | 3 (37.5) |  |
| **History of Contraception Use<sup>&</sup>** |  |  |  | 1.00 |
| *Hormonal Contraception<sup>#</sup>* | 5 (31.2) | 2 (25.0) | 3 (37.5) |  |
| *None* | 1 (6.2) | 1 (12.5) | 0 (0.0) |  |

After filtering, taxonomic classification of the vaginal specimens identified 20 vaginal microbial taxa, listed in order of relative abundance: *Lactobacillus iners*, *Lactobacillus crispatus*, *Lactobacillus jensenii*, *Gardnerella, Lactobacillus gasseri, Prevotella,* unclassified *Lactobacillus* spp.*, Streptococcus, Aerococcus, Sneathia, Dialister,* BV-associated bacterium 1 (BVAB1; also known as *Candidatus Lachnocurva vaginae*)^41^*, Megasphaera, Fannyhessea* (formerly classified as *Atopobium*)*, Fastidiosipila, Mobiluncus, Parvimonas, Limosilactobacillus, Lactobacillus intestinalis*, and *Ligilactobacillus*.

Of the 420 vaginal specimens included in this analysis, 137 were pre-iBV specimens from eight iBV cases and 283 were specimens from eight matched healthy controls. Our final trained model was designed to predict whether a participant would develop iBV within 14 days of collection (pre-iBV) or remain healthy using a single vaginal specimen.

Our workflow for obtaining specimens, generating data, and training models is shown in **Figure 1**.

### Predictive Modeling for Early Detection of iBV

To assess if characterization of the vaginal microbiome could aid in early detection of iBV we initially trained ANN models using the IAA of 20 identified vaginal taxa in our cohort. Following parameter and architecture optimizations, the final model achieved a peak accuracy of 100% on training data and 97.7% on testing data. In addition to attaining high accuracy, the 20-feature model also achieved high sensitivity and specificity, at 96.1% and 98.2%, respectively (**Figure 2**, **Table 2, Figure S1**).

**Figure 2.**
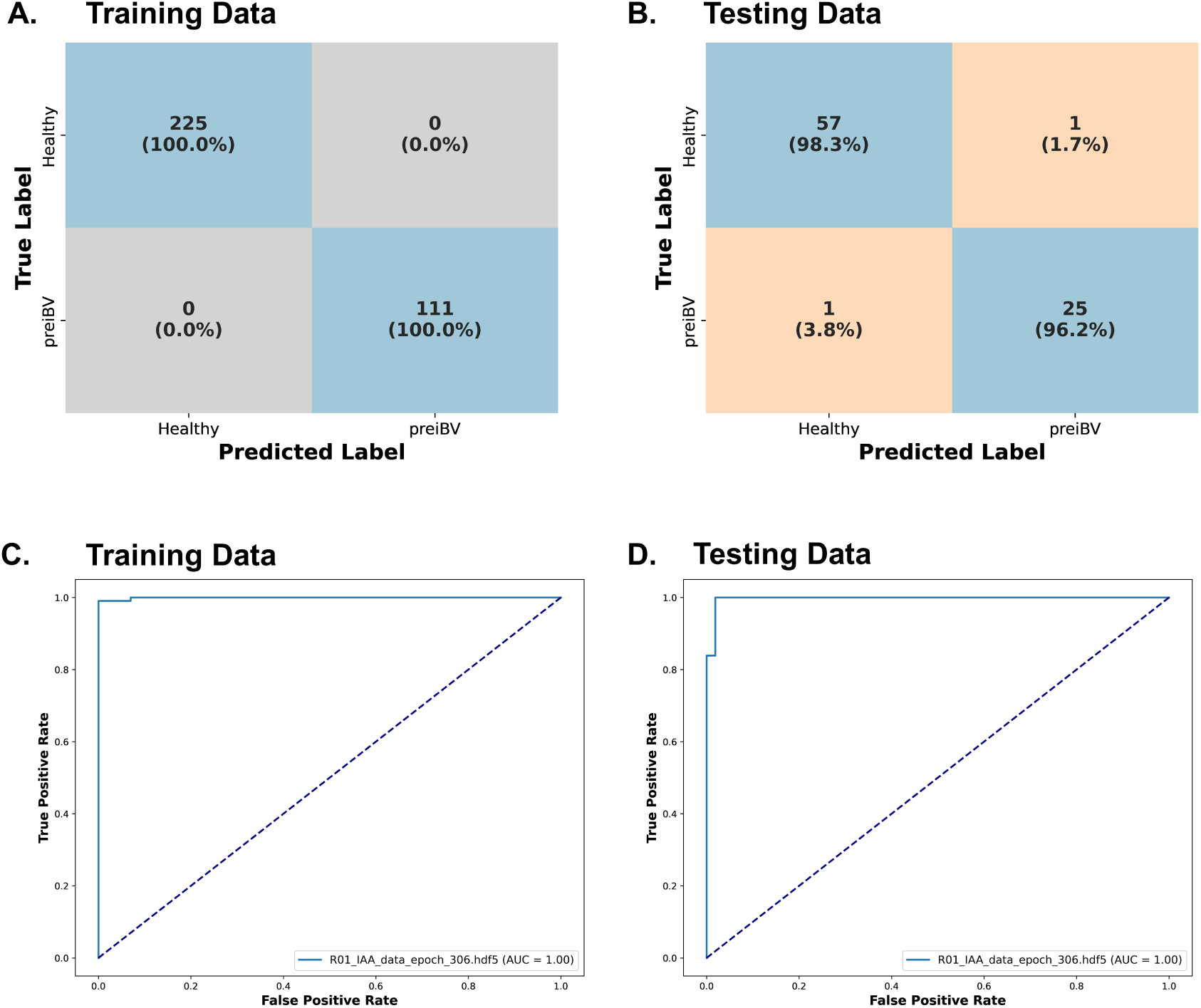
20-Feature Model Classification Performance & AUC. Summary of ANN modeling performance trained using inferred absolute abundance (IAA) of 20 common vaginal taxa (n = 420). (A-B) Confusion Matrices model classifications in relation to true classification of (A) training data (n = 336) and (B) testing data (n = 84). Tiles containing correct classifications are shown in blue and tiles containing incorrect classification are shown in orange. Tile labels specify the number of specimens in each category and percentage of specimens in each category based on their true classification. (C-D) ROC curve of ANN model performance on (C) training data and (D) testing data

**Table 2.** 20-Feature Model Performance Metrics. Metrics calculated from model classification accuracy on training data (n = 336) and testing data (n = 86) Abbreviations: ROC-AUC = Receiver Operating Characteristic – Area Under the Curve, PPV = Positive Predictive Value, NPV = Negative Predictive Value

| **Metric** | **Train** | **Test** |
| --- | --- | --- |
| **Accuracy** | 100% | 97% |
| **Sensitivity** | 100% | 96% |
| **Specificity** | 100% | 98% |
| **F1 Score** | 100% | 96% |
| **ROC-AUC** | 100% | 99% |
| **PPV** | 100% | 96% |
| **NPV** | 100% | 98% |
| **Cohen Kappa** | 100% | 94% |

**Table 3.**
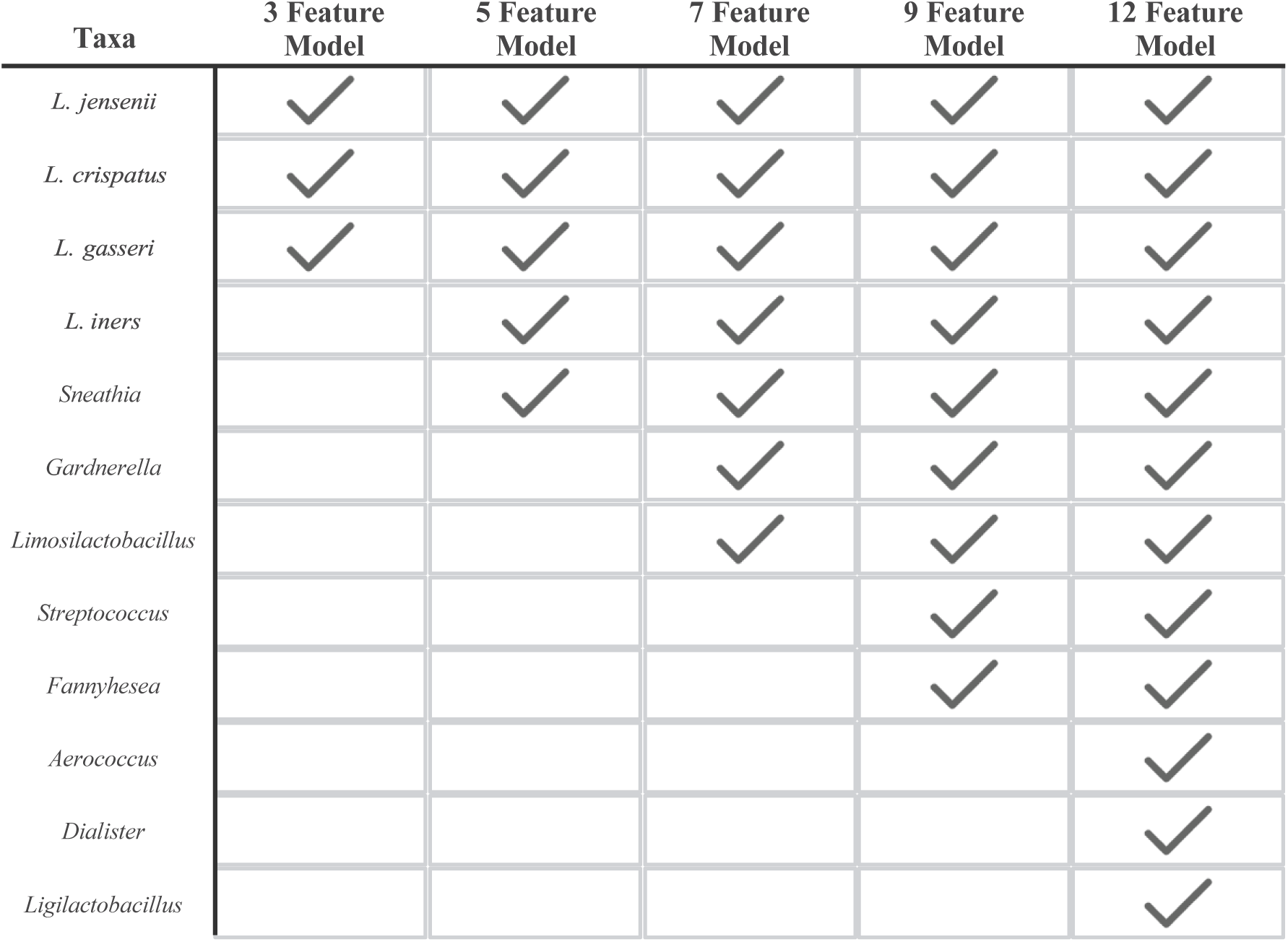
Taxa included as features in each model subset. Checkmark indicates IAA of the corresponding taxa included as features in the specified subset model.

### Role of Microbial Features in Pre-iBV Classification

SHapley Additive exPlanations (SHAP) analysis of the 20-feature model assessed how each taxa (feature) contributed to model predictions (**Figure 3)**.^40^ This analysis revealed that several vaginal *Lactobacillus* spp. ranked as the most important features for model accuracy.^42^ Specifically, high IAAs of *L. crispatus* and *L. jensenii* was associated with classifying specimens as healthy, while low IAA of *L. iners* trended towards healthy classifications. Interestingly, a low IAA of *L. gasseri* contributed to classifying specimens as healthy, whereas a high IAA of *L. gasseri* associated with pre-iBV classifications. A higher abundance of BV-associated bacteria such as *Gardnerella*, *Sneathia*, BVAB1, *Fannyhessea,* and *Aerococcus* were found to be associated with pre-iBV predictions.

**Figure 3.**
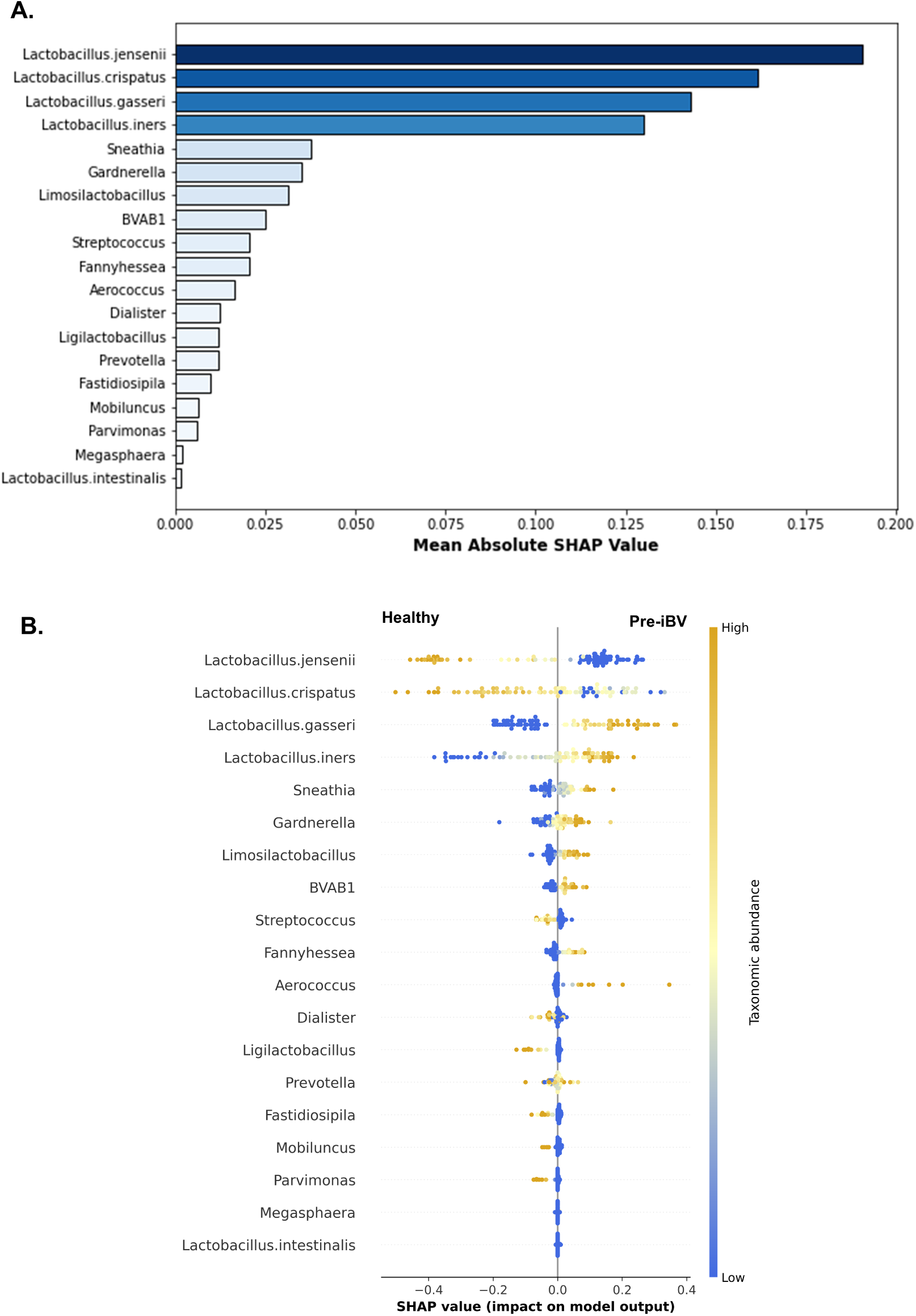
Feature importance and Feature Use Determined from SHAP analysis. (A) Importance of features included in the 20-Feature model determined by Mean Absolute SHAP value of features in the model testing data (n = 84). (B) Beeswarm plot indicates how features contribute to model predictions for each specimen in the model testing data (n = 84). IAA abundance of each taxa is reflected by color gradient, orange indicates high IAA abundance and blue indicates low IAA abundance relative to other specimens. X-axis position indicates whether the feature contributes to predicting a specimen is classified as Healthy (Left) or preiBV (Right). Distance from O on the X-axis indicates how much each feature contributes to accurately classifying an individual specimen.

### Minimum Feature Analysis

To evaluate the number of vaginal taxa required to accurately classify specimens, successive models were constructed with training a strategy using a limited subset of important features. Specifically, five subset models were generated though training using the top three, five, seven, nine, and twelve most important features, respectively (**Figure 4**, **Figure S2, Table S2**). The model trained on the top three most important features, (*L. jensenii, L. crispatus*, and *L. gasseri*) had a peak training accuracy of 90.1% and a testing accuracy of 94.0%. In training and testing data, the three-feature model had decreased sensitivity (training = 78.3%, testing = 84.6%) compared to specificity (training = 96.0%, testing = 98.2%), indicating this model classified pre-iBV specimens less accurately than healthy specimens. When trained using the top seven, nine, and twelve features, models achieved peak training accuracies ≥ 98%, as well as improved sensitivity (sensitivity > 96%), demonstrating improved performance with the inclusion of additional taxa as well as improved classification of pre-IBV specimens. These findings indicate that while surveying a few key vaginal taxa is sufficient to classify specimens as pre-iBV or healthy, incorporating additional taxa enhances the classification of pre-iBV specimens.

**Figure 4.**
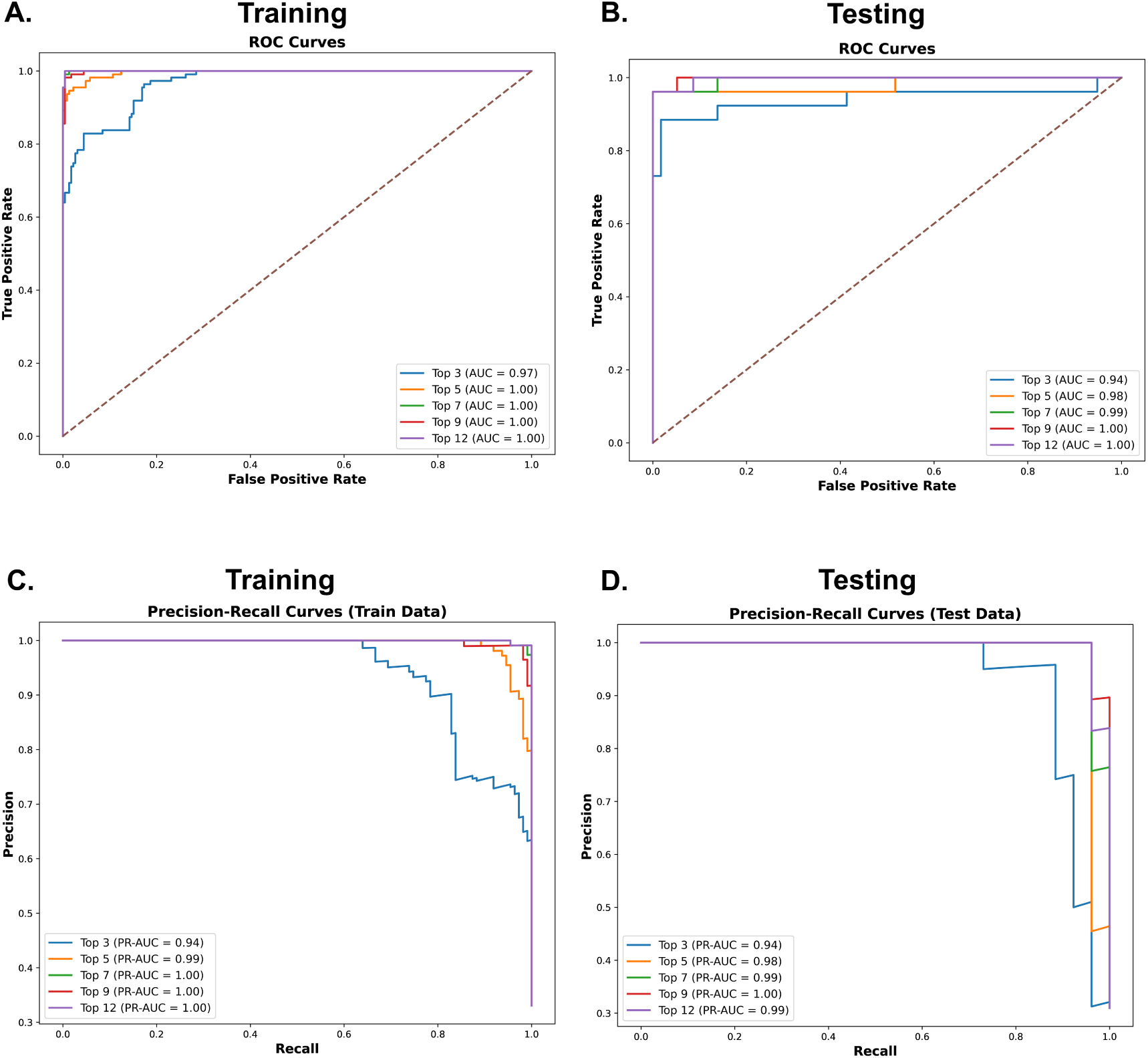
N-Feature Models Receiver Operating Curves and Precision-Recall Curves. Performance of models trained on the three, five, seven, nine, and twelve most important features determined by feature mean absolute SHAP value are visualized as (A-B) receiver operating curves and (C-D) Precision-Recall Curves. Curves calculated from training data (n = 336)are shown in (A,C)and curves calculated from testing data (n = 84) are shown in (B,D).

SHAP analysis was applied to determine how features were utilized by the n-feature models (n = top three, five, seven, nine, and twelve features, **Figure S3**). Across all n-feature models, *Lactobacillus* spp. consistently emerged as the most important taxa, similar to findings from the 20-feature model. In subset models where *Gardnerella* was included as a feature, *Gardnerella* ranked as the fourth most important feature, following *Lactobacillus* spp. Similar to the 20-feature model, subset models, high IAAs of *Gardnerella*, BVAB1, and *Sneathia* contributed to classifying specimens as pre-iBV, while lower IAA levels of these taxa favored classification as healthy. Results from SHAP analysis reflect *Lactobacillus* spp. and BV-associated genera such as *Gardnerella*, BVAB1, and *Sneathia*, impacting model predictions across the feature sets tested.

### Race-Specific Models: Performance and Feature Utilization

Considering there is evidence suggesting that the composition of the vaginal microbiota differs based on race,^18,19^ we trained models separately using specimens from black (B-Model) (total n = 176, training n = 140, testing n = 36) and white (W-Model) (total n = 234, training n = 187, testing n = 47) participants.^18^ After training, both the B-Model and the W-Model reached 100% accuracy on both the training and testing datasets (**Figure 5, Table S2,** and **Figure S4)**.

**Figure 5.**
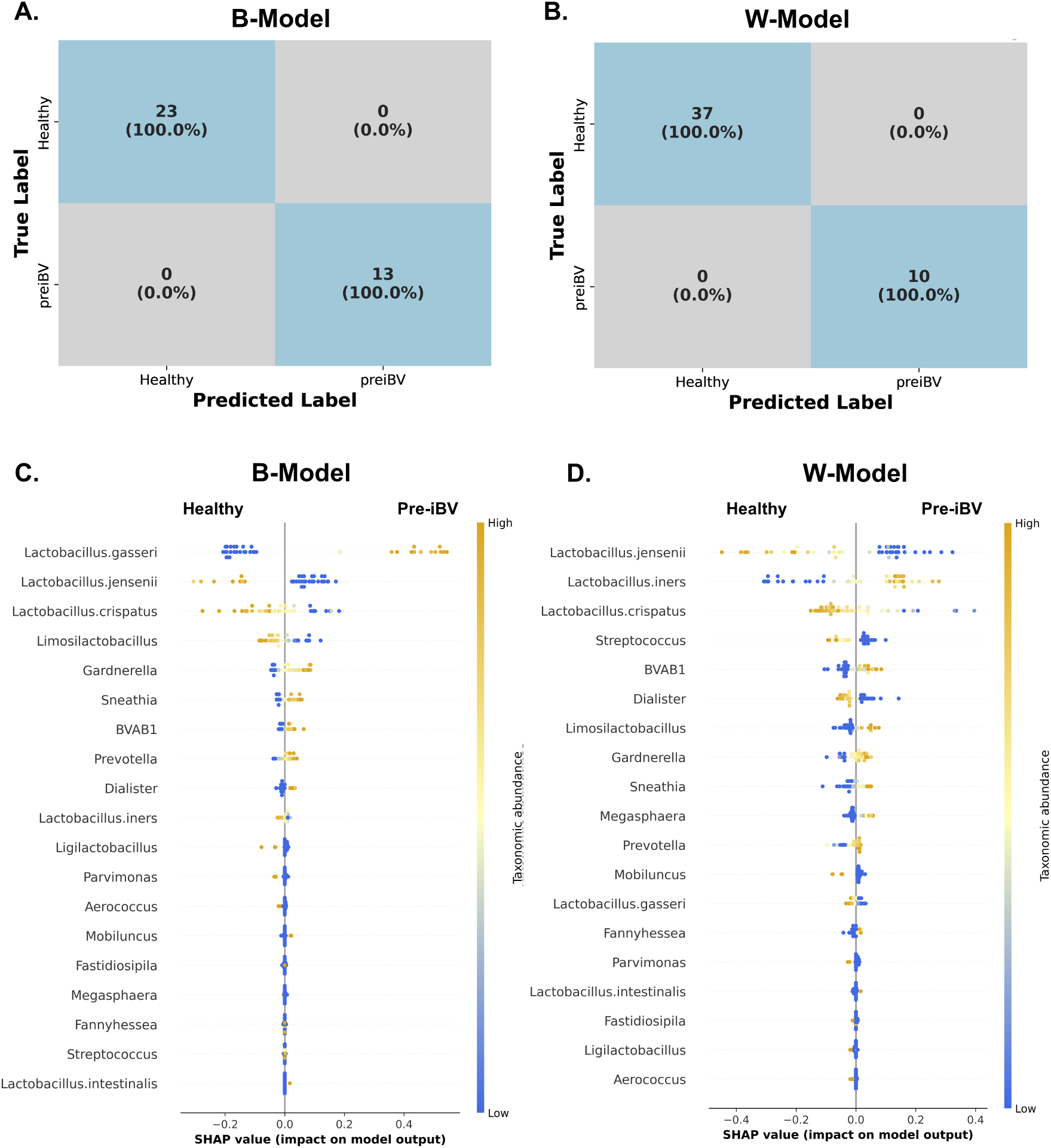
Classification performance and SHAP analysis of Models trained on Race­ specific data. (A-B) Confusion matrices of classification results of testing data on models trained on specimens from (A) black participant specimens (n = 176) or (B)white participant specimens (n = 234). Blue tiles indicate correct classifications by the model. Tile labels specify the number of specimens in each category and percentage of specimens in each category based on theirtrue classification. (C-D) Beeswarm plots indicate feature importance and how features contribute to model predictions. SHAP analysis was performed on testing data from models trained using specimens from (C) black participants (n = 36 testing specimens) or (D) white participants (n = 47 testing specimens). IAA abundance of each taxa is reflected by color gradient, orange indicates high IAA abundance and blue indicates low IAA abundance relative to other specimens. X-axis position indicates whether the feature contributes to predicting a specimen is classified as healthy (Left) or preiBV (Right). Distance from O on the x-axis indicates how much each feature contributes to accurately classifying an individual specimen.

In both models, *Lactobacillus* spp. ranked as the top three most important features, however, the specific species and how the species contributed to model predictions differed between the two models. In the B-Model, *L. gasseri* was the most important feature with a high IAA being predictive of a pre-iBV state, although ranked as the 13^th^ most important feature in the W-model. On the other hand, in the W-Model and B-Model, *L. jensenii* ranked as second the most important feature. Additionally, *L*. *iners* ranked as the second most important feature in the W-model but was minimally important to predictions in the B-model, ranking as the 10th most important feature. In both models, a high relative IAA of BV-associated bacteria including *Prevotella*, BVAB1, *Gardnerella*, and *Sneathia* were found to be predictive of pre-iBV, and a low relative IAA of these taxa was predictive of healthy vaginal microbiota. These findings indicate that, while both models were highly accurate, the role of *L. iners* and *L. gasseri* in the onset of iBV may differ depending on a person’s race. This highlights potential race-specific differences in the onset of iBV.

To determine the minimum number of features required to accurately classify a specimen as pre-iBV or healthy for race-specific models, similarly, models were trained using the top three, five, seven, nine, and twelve most important features (**Figure S5 & S6**). The specific taxa included in race-specific N-feature models are listed in **Table S3 & S4**. Both the three-feature B and W models were highly accurate. The three-feature model trained on data from black participants accurately categorized 96.4% of black participants in training data and 97.2% in testing data. Similarly, the three-feature model trained on white participants accurately classified 98.3% of white participants in the training data and 100% in the testing data (**Figure 6**). Race-specific models achieved 100% accuracy in training and testing data when trained on a minimum of five features in black participants and seven in white participants (**Table S5 & S6**), suggesting that applying race-specific models improves detection of pre-iBV and requires fewer taxa for accurate classification compared to models trained on the complete cohort.

**Figure 6.**
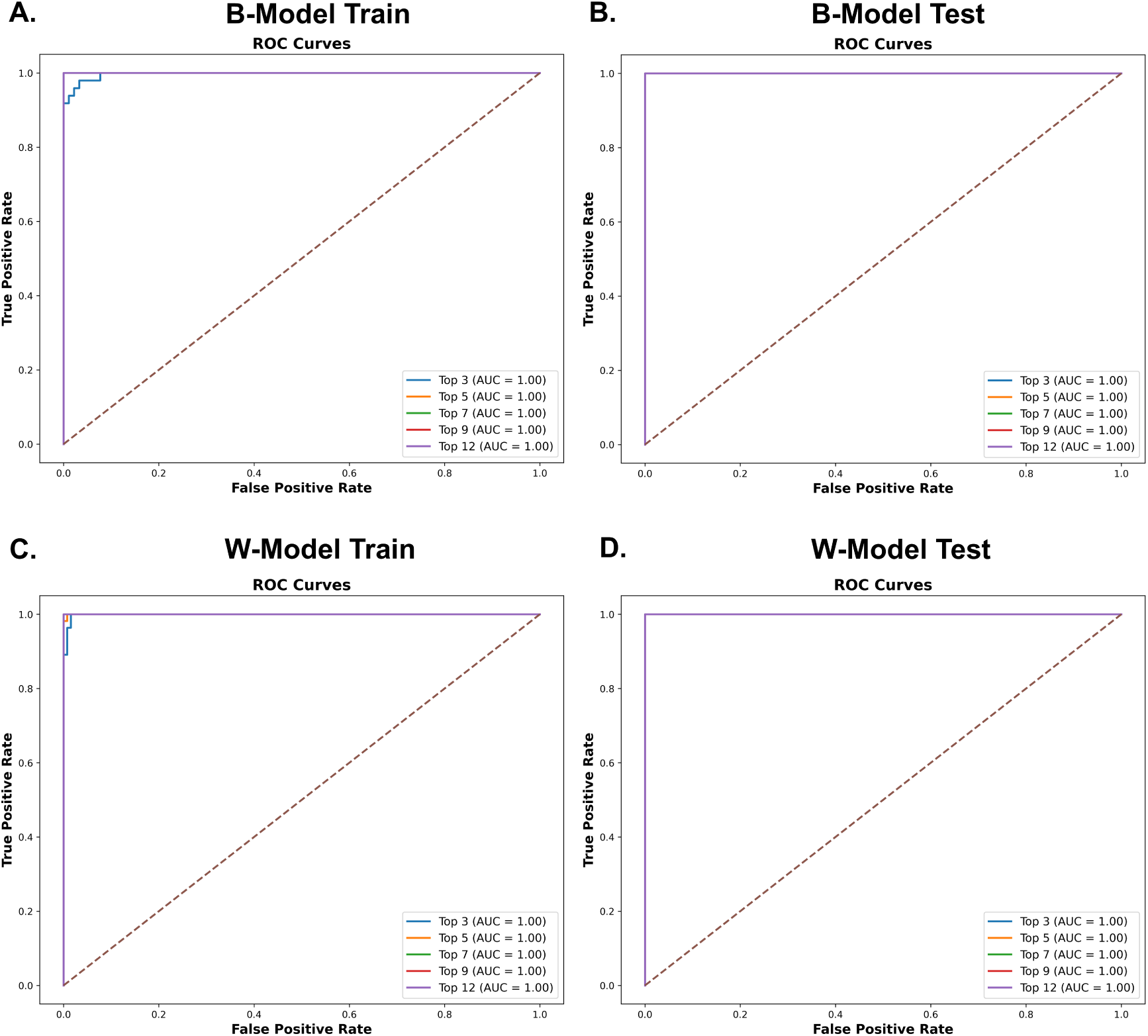
Receiver Operating Curves of Race Specific N-Feature Models. (A-B) Receiver Operating Curves of models trained the top three, five, seven, nine, twelve most important features to B-model predictions based on classification on (A) training data (n = 140) and (B) testing data (n = 36). (C-D) Receiver Operating Curves of models trained the top three, five, seven, nine, and twelve most important features to W-model predictions based on classification on (C) training data (n =187) and (D) testing data (n = 47).

To further assess how the models used each feature, SHAP analysis was performed on each race-specific n-feature model (**Figure S7 & S8**). Across all models, independent of race or feature subset, *Lactobacillus* spp. consistently ranked among the most important features for model predictions. In both black and white participant n-feature models, a high IAA of *L. crispatus* and *L. jensenii* were associated with a healthy classification, similar to what was observed in the 20-feature models. Also, *L. gasseri* was the most important feature for classifying specimens in black participants across all model subsets, but it ranked as one of the least important features in n-feature models trained and tested on white participants. In all models, a high relative IAA of common BV-associated bacteria including *Gardnerella*, *Sneathia*, and BVAB1 was associated with classifying specimens as pre-iBV. These findings demonstrate training and testing models separately on black and white participants allows for the identification of race-specific patterns, which likely contributed to enhanced classification accuracy.

## DISCUSSION

In this study, we developed an ANN model capable of accurately predicting the onset of iBV up to 14 days in advance from a single vaginal specimen. Our models relied on 420 individual specimens, enabling fine resolution of microbiome changes associated with iBV onset. The model achieved high accuracy, sensitivity, and specificity using IAA of vaginal taxa to make predictions.^29^ This predictive capability enables early detection of iBV, allowing for proactive interventions to potentially prevent its development.

By analyzing our model, to understand how it accurately classified specimens, microbial patterns correlating with pre-iBV states were revealed. From SHAP analysis, we found common vaginal *Lactobacillus* spp. were the most important taxa to accurately classify specimens as healthy or pre-iBV. As expected, a higher abundance of *L. crispatus* and *L*. *jensenii* were indicative of stable vaginal communities while a higher abundance of BV-associated bacterial genera such as *Gardnerella*, *Sneathia,* and BVAB1 were indicative of pre-iBV communities.^18,27,42^ Somewhat to our surprise, *L. gasseri* abundance positively associated with pre-iBV predictions, although prior studies have shown *L. gasseri* strains produce less L-lactic acid and hydrogen peroxide, compared to *L. crispatus.*^43,44^ The relationship between *L. gasseri* and iBV is incompletely understood due to prior work primarily relying on compositional approaches to quantify vaginal microbiota.^42^ While SHAP analysis assesses correlative relationships between features and predictions, our findings suggest additional emphasis should be placed on further assessing the role of *L. gasseri* in iBV, using non-compositional methods to measure *L. gasseri* abundance (i.e., qPCR/IAA). Furthermore, our results support the notion that *L. crispatus* is critical to iBV protection, while less common *Lactobacillus* spp. such as *L. gasseri* and *L. jensenii* may vary in their protective capacity due to strain variation and host features, including an individual’s racial background.

Minimum feature analysis revealed that only a few taxa were necessary for highly accurate predictions. Models trained on the top five features (*L. jensenii, L. crispatus, L. gasseri, L. iners*, and *Sneathia*) achieved >95% accuracy. While additional vaginal taxa improved accuracy, the strong performance of minimal models suggests that simplified approaches to characterizing the vaginal microbiome may be sufficient for specific research and diagnostic purposes. For example, using targeted qPCR panels of five to seven organisms could be feasible alternatives to complex sequencing-based profiling for early iBV detection.

Models developed from race-specific subgroups improved model accuracy and revealed distinct microbial signatures contributing to pre-iBV predictions in black and white participants*. L. gasseri* was found to be highly important to pre-iBV predictions in models trained on the black subgroup, although this was not reflected in models trained on the white subgroup. Additionally, *L. iners* was a key predictor of pre-iBV in white-specific models but ranked low in importance for predicting pre-iBV in black participants. These findings support prior work demonstrating that changes in the composition of vaginal bacterial communities over time differs by race.^18^ The implications of these race-specific differences are profound and suggest that personalized diagnostic approaches may be necessary for the early detection of iBV. In future work, we hope to validate these findings using larger and more diverse cohorts.

Early detection of iBV could have significant clinical implications, allowing for early clinical interventions such as live biotherapeutics, prophylactic antibiotics, and/or behavioral modifications. Furthermore, high accuracy observed on race-specific models underscores the importance of considering demographic and biological diversity in developing microbial models and potentially in clinical practice.

While we have rigorously conducted our analysis and added valuable insight into the early detection of iBV, there are several limitations to our study. Although our longitudinal data were rich, our cohort was relatively small. Additionally, the cohort used to train our ANN models reflects the racial demographics of Birmingham, AL, as it was primarily composed of participants that identified as black or white, limiting our ability to identify specific racial/ethnic relationships between pre-iBV and the vaginal microbiome for other groups such as Hispanics, Asians, and Native Americans.^45–47^ Future studies externally validating our models with larger cohorts from more racially diverse studies will be required to verify model validity. Furthermore, since SHAP analysis only identifies correlations between model predictions and microbial taxa, future studies will be necessary to validate the relationships between vaginal microbiota and pre-iBV classifications. Similarly, future studies will be required to identify the underlying biological mechanism(s) mediating the microbial relationships found through SHAP analysis.^48^ Although our model was trained on a relatively small cohort with specific enrollment criteria, the high accuracy achieved reflects the strong biological patterns underlying the pre-iBV state.

In summary, this study demonstrates that machine-learning approaches are highly effective in predicting iBV prior to its onset, which has the potential to be applied to the preemptive diagnosis of iBV from a single vaginal specimen. Our findings emphasize the role *Lactobacillus* spp. play in regulating vaginal microbiome dynamics as well as race-specific differences in microbial predictors of pre-iBV states. Early detection of iBV could lead to wider adoption of clinical interventions useful in the prevention of iBV such as live biotherapeutics, prophylactic antibiotics, and/or behavioral modifications.^49,50^ Integrating personalized microbiome-based diagnostics capable of early detection of vaginal infections could revolutionize the management of BV and other important vaginal health conditions.

## Data Availability

The sequencing data used for the purposes of this study has been made available through the NCBI Sequence Read Archive (SRA) BioProject accession (PRJNA1232196). All other data produced in the present study are available upon reasonable request to the authors.

https://www.ncbi.nlm.nih.gov/bioproject/?term=(PRJNA1232196)%20AND%20bioproject_sra[filter]%20NOT%20bioproject_gap[filter]

## Data Sharing Statement

The sequencing data used for the purposes of this study has been made available through the NCBI Sequence Read Archive (SRA) BioProject accession (PRJNA1232196).

## Declaration of Interests

PŁ reports grants from the National Institutes of Health (NIH), National Cancer Institute (NCI) during the conduct of the study. MMHK reports grants from the NIH and NCI during the conduct of the study, as well as consulting fees from Freya Biosciences outside the submitted work. NC reports financial support by the Portuguese Foundation for Science and Technology. CAM reports grants to her institution from NIH, National Institute of Allergy and Infectious Diseases (NIAID), Gilead, BioNTech, and Abbott; consulting fees from Abbott, BioNTech, bioMérieux, and Cepheid; honorarium from Abbott, Elsevier, and the Merck Manuals; and royalties from UpToDate. CMT reports grants to his institution from NIH and reports consulting fees from Tulane University. No disclosures are reported by the other authors.

## Acknowledgements

This work was supported by the National Institute of Allergy and Infectious Diseases (grants R01AI146065 to CAM and R01AI118860 to AJQ and CMT), National Center for Advancing Translational Sciences (grant UL1TR003096 to the University of Alabama-Birmingham Center for Clinical and Translational Science), National Cancer Institute (grant U54CA143924 to MHK), and the National Science Foundation (grant 2018936 to CMT). The funders were not involved in study design, data collection, model design, data analysis, data interpretation, or manuscript drafting. A U.S. Provisional Patent Application No. 63/7789,89 have been filed by LSUHSC regarding this work. This application is entitled “Predicting Bacterial Vaginosis Development Using Artificial Neural Networks.” This work was presented as oral presentation #028 at the 2024 Infectious Diseases Society for Obstetrics and Gynecology in Portland, OR, on August 1-3, 2024.

## Author Contributions

JE, JL, NC, CAM, and CMT conceived the study and were in charge of overall direction and planning. JE designed the initial model and the computational framework. Additional model development, data management, and analysis was performed by JE, JL, and KJA. JE, CA, CJ, and ML performed the molecular experiments. KJA and KJG clinically categorized patient specimens and established the specimen bank. JE and JL wrote the manuscript with input from all authors. MHK, PL, NC, AJQ, CAM, and CMT jointly supervised the study and provided oversight during the manuscript editing process. All authors discussed the results, contributed to the final manuscript, and approved the submitted version.

## Notes

### Competing Interest Statement

CMT has received research grant support to his institution from NIH and reports consulting fees from Tulane University. CAM has received research grant support to her institution from NIH/NIAID, Gilead, BioNTech, and Abbott; and reports consulting fees from Abbott, BioNTech, bioMerieux, and Cepheid; honorarium from Abbott, Roche, Elsevier, and the Merck Manuals; and royalties from UpToDate. NC reports financial support was provided by the Portuguese Foundation for Science and Technology. MMHK reports grants from the NIH and NCI during the conduct of the study, as well as consulting fees from Freya Biosciences outside the submitted work. PŁ reports grants from the National Institutes of Health NIH, NCI during the conduct of the study. No disclosures were reported by the other authors.

### Clinical Protocols

https://pubmed.ncbi.nlm.nih.gov/38316599/

### Author Declarations

This study was approved by the University of Alabama at Birmingham Institutional Review Board, Protocol #300004547, with a Reliance Agreement at LSUHSC New Orleans.

